# Automated Sleep Stage and Event Detection Algorithms Using Quality-Controlled PSG Annotations

**DOI:** 10.64898/2025.12.15.25342334

**Authors:** Michiru Kaneda, Sho Ogaki, Tomoyuki Nohara, Syuhei Fujita, Naoshi Osako, Tomoko Yagi, Yasuhiro Tomita, Takanori Ogata

**Affiliations:** ACCELStars, Inc., 7-3-1 Hongo, Bunkyo-ku, 113-0033, Tokyo, Japan; School of Medical Technology, Kurume University, 67 Asahi-machi, Kurume, 830-0011, Fukuoka, Japan; Sleep and Respiratory Medicine, Toranomon Hospital, 2-2-2 Toranomon, Minato-ku, 105-8470, Tokyo, Japan; Graduate School of Medicine, The University of Tokyo, 7-3-1, Hongo, Bunkyo-ku, 113-0033, Tokyo, Japan; Faculty of Medicine, Juntendo University, 2-1-1 Hongo, Bunkyo-ku, 113-8421, Tokyo, Japan

**Keywords:** Polysomnography, Sleep Stage Classification, Arousal Detection, Respiratory Event Detection, Inter-Scorer Agreement, Automated Sleep Analysis

## Abstract

**Study Objectives:** To develop machine-learning models for sleep stage classification, arousal detection, and respiratory event detection from overnight polysomnography, and to evaluate their performance relative to expert scorers.

**Methods:** Overnight polysomnography recordings were obtained from healthy participants and participants referred for suspected sleep-disordered breathing. Four certified scorers completed calibration sessions and generated reference annotations for sleep stages, arousals, and respiratory events. A subset of recordings was independently annotated by all scorers to support consensus analyses, enabling direct comparison between model outputs and human inter-scorer agreement. Gradient-boosted decision tree models were trained using hand-crafted features derived from standard physiological signals.

**Results:** Sleep stage classification achieved accuracy 0.840, Cohen’s kappa 0.791, and F_1_-score 0.841, with limits of agreement for total sleep time of approximately *±*0.5 h. Arousal detection achieved an F_1_-score of 0.733, with limits of agreement for the arousal index of approximately *±*15 events/h. Respiratory event detection achieved an F_1_-score of 0.818, with limits of agreement for the apnea–hypopnea index also within approximately *±*15 events/h. In consensus analyses, model performance was comparable to human inter-scorer agreement for sleep stages and arousals, while remaining below human inter-scorer agreement for respiratory events, despite high absolute performance relative to prior studies.

**Conclusions:** The proposed models achieved performance approaching human-level agreement across major sleep scoring tasks. These findings indicate that high consistency in expert annotations is a key factor underlying robust model performance and support the use of quality-controlled annotations for developing reliable automated sleep analysis systems.

**Statement of significance:** Manual scoring of overnight sleep studies remains a major bottleneck in sleep medicine, limiting efficiency, consistency, and large-scale research. This study demonstrates that interpretable automated analysis can achieve performance approaching human-level agreement for core sleep scoring tasks when reference annotations are highly consistent. By directly comparing model outputs with calibrated inter-scorer agreement, the results show that annotation quality is a key determinant of attainable accuracy, rather than model complexity alone. Such systems may provide stable and reproducible reference outputs that support clinical decision making, scorer training, and standardization across centers. Important remaining challenges include validation across institutions and populations, robustness to real-world signal artifacts, and extension to clinically meaningful subtypes of respiratory events.

## Introduction

Sleep disorders such as insomnia and sleep-disordered breathing (SDB) are highly prevalent conditions associated with increased cardiovascular, metabolic, and neurocognitive risks [1–4]. Accurate evaluation of sleep architecture and respiration is therefore essential for diagnosis and management, with polysomnography (PSG) remaining the gold standard modality [5, 6]. By capturing comprehensive physiological signals, PSG enables precise assessment of sleep physiology and disease severity, supporting optimal clinical and therapeutic decision making [7].

PSG typically records multiple physiological channels, including electro-encephalography (EEG), electrooculography (EOG), electromyography (EMG), electrocardiography (ECG), respiratory effort, airflow, and oxygen saturation. From these data, trained scorers manually annotate events such as sleep stages, arousals, and respiratory abnormalities [8]. The American Academy of Sleep Medicine (AASM) manual provides standardized scoring rules that are widely adopted across sleep laboratories [9–11].

Although these criteria have improved standardization, PSG scoring remains predominantly based on visual inspection by trained scorers. This process is labor intensive, time consuming, and dependent on specialized expertise, often limiting diagnostic throughput. Because PSG interpretation inherently depends on human judgment, scoring accuracy is constrained by perceptual and cognitive limitations, resulting in notable inter-scorer variability even under AASM guidelines [12–16]. Variations in institutional practices, scorer training, and interpretation further challenge standardization, limiting both clinical consistency and research reproducibility. Consequently, there has been increasing interest in automated PSG analysis to streamline clinical workflows, reduce scoring burden, and improve inter-scorer reliability.

Earlier studies employed feature-based machine learning approaches such as support vector machines, random forests, and decision trees for automatic sleep stage classification [17–22]. Subsequent works introduced deep learning architectures capable of learning temporal and spectral representations directly from raw data, achieving improved accuracy [23–27]. More recently, interpretable modeling approaches have emerged, including image-based representations and cross-modal transformer architectures designed to provide human aligned explanations of PSG predictions [28–30]. At the same time, traditional machine learning pipelines have been shown to perform comparably to state of the art deep learning models [31].

Similar progress has occurred in automated detection of arousals and respiratory events. Early automated approaches relied on handcrafted features derived from EEG, ECG, airflow, and respiratory effort [32–35], whereas later research increasingly adopted deep learning models that directly capture temporal dependencies and event durations from PSG signals [36–40].

Furthermore, a recent hybrid pipeline integrates deep learning and signal processing to analyze sleep staging, arousal, and respiratory events, and additionally detects limb movements within the same framework [41].

Many of these studies rely on publicly available PSG databases, which provide valuable benchmarks for cross study comparison. However, these datasets differ considerably in their scoring conventions and preprocessing standards. Older databases were annotated according to the Rechtschaffen and Kales (R&K) criteria [42], and although conversion guidelines to the AASM system exist [43], they are not perfectly equivalent. Moreover, studies vary in how they select and segment data—such as the definition of lights out and lights on intervals, the treatment of high noise segments, and the allocation of data into training and test sets—as well as in how they match events for evaluation. Because these methodological details are not standardized across datasets or studies, direct numerical comparison of reported performance metrics is inherently difficult.

In addition to these inconsistencies, the reliability of human generated annotations themselves remains uncertain. Several studies have investigated uncertainty in sleep stage scoring and compared the performance of automated systems with that of human scorers [41, 44, 45]. These studies have demonstrated that substantial uncertainty exists in manual scoring, and that automated models can achieve levels of agreement comparable to trained experts and, in some cases, exceed the consistency of individual scorers relative to a selected reference standard.

Importantly, recent work has emphasized that disagreement among human scorers often reflects intrinsic ambiguity in sleep stage transitions rather than annotation errors alone. In this context, probabilistic sleep staging frameworks have been proposed to characterize such uncertainty by assigning a distribution over sleep stages to each epoch, thereby providing a quantitative description of scorer disagreement and transitional physiology beyond conventional hard labels [46–48]. Collectively, these findings indicate that both human agreement and apparent model performance are constrained by the inherent uncertainty of PSG scoring, and that improvements in automated analysis should be interpreted in relation to annotation reliability and ambiguity.

In the present study, a high-quality reference dataset was established through a structured calibration process rather than through adjudicated consensus labeling. Multiple certified sleep scorers first participated in targeted calibration sessions to harmonize their scoring criteria for sleep stages, arousals, and respiratory events. After calibration, each scorer independently annotated PSG recordings under unified scoring guidelines, yielding a set of consistent and quality-controlled labels suitable for algorithm development. A subset of recordings was double scored to quantify inter-scorer reliability, which was then compared with model performance to assess the extent to which the automated system approximated human level agreement. This design avoids the variability introduced by uncalibrated datasets while providing a set of consistently generated, high-quality annotations for model development and evaluation.

Using this rigorously standardized annotation set, a gradient-boosted decision tree (GBDT) [49]-based framework was developed and evaluated for sleep stage classification, arousal detection, and respiratory event detection. Model performance was assessed relative to multiple expert scorers to provide a benchmark grounded in high-quality, consistently generated PSG annotations.

## Methods

### Ethics Statements

This study was approved by the Ethics Committee of Medical Corporation Kouseikai Sone Clinic (Protocol IDs: SUG08869 and SUG08884) and by the Ethics Committee of Koga Hospital 21 (Protocol ID: 21-033). Written informed consent was obtained from all participants in the prospective studies at Sone Clinic. For the retrospective study at Koga Hospital 21, the requirement for written informed consent was addressed through an opt-out process implemented by the hospital and disclosed on its website, in accordance with the approved protocol. All data were anonymized prior to analysis, and no identifying information is included in this manuscript.

### Dataset

Three types of overnight PSG datasets were used in this study. The first dataset consisted of PSG recordings acquired from healthy participants in a dedicated in-house sleep research laboratory equipped for controlled overnight monitoring, with SOMNO BT Plus (SOMNOmedics AG) devices. The second and third datasets consisted of PSG data obtained at two different collaborating clinical sleep centers from participants referred for evaluation of suspected SDB, primarily obstructive sleep apnea (OSA). Alice 6 LDx (Royal Philips) and SOMNOtouch RESP (SOMNOmedics AG) devices were used to acquire PSG data at the two clinical centers, respectively. All PSG recordings were obtained using standard clinical PSG montages in accordance with the AASM Scoring Manual.

For the first and second datasets, six EEG channels recommended by the AASM manual for sleep staging were included: F3–M2, F4–M1, C3–M2, C4–M1, O1–M2, and O2–M1. The third dataset included only three EEG channels, F3–M2, C3–M2, and O1–A2. All datasets included two EOG channels, E1–M2 and E2–M1 and one chin EMG channel, as well as one nasal pressure airflow channel and one SpO_2_ channel. Additional clinical PSG channels were also recorded as part of a standard clinical PSG montage, including thermistor airflow, thoracoabdominal effort signals, and bilateral anterior tibialis EMG. These channels were available during manual scoring, but only the channels used for feature extraction are described here in detail.

Although the third dataset has only three EEG channels, it is enough to perform manual analysis if all three channels are clear. On the other hand, this study focuses on the development with full six EEG channels, and the third dataset was used only for scorer calibration, not for model development or evaluation.

In the following tables, the term “Healthy” refers to the first dataset of PSG recordings from healthy participants, while “SDB” refers to the second dataset of PSG recordings from participants with suspected sleep-disordered breathing. The third dataset, which was used only for scorer calibration, is referred to as “Calibration SDB”.

Table 1 summarizes the sampling frequencies of the PSG channels from which features were extracted.

**Table 1.** Sampling frequencies (Hz) of PSG channels.

| Channel | SOMNO BT Plus | Alice 6 LDx | SOMNOtouch RESP |
| --- | --- | --- | --- |
| Electroencephalography | 256 | 500 | 256 |
| Electrooculography | 256 | 500 | 256 |
| Electromyography | 256 | 500 | 256 |
| Nasal pressure airflow | 256 | 100 | 256 |
| SpO<sub>2</sub> | 4 | 1 | 4 |

For the model development and evaluation, a total of 604 recordings from healthy participants and 245 recordings from participants with suspected SDB were initially obtained as the first and second datasets, respectively. Recordings were screened to include only complete overnight sessions with sufficient signal quality and no major signal loss. For participants with multiple recorded nights, a single night was selected to avoid duplication at the participant level. After this quality control, 473 healthy recordings and 244 SDB recordings were included in the sleep stage classification study (Table 2), as these recordings were sufficiently clear to allow full night sleep stage classification. From this quality-controlled subset, 412 healthy recordings and 178 SDB recordings were further included in the arousal detection study, reflecting the additional requirement that data quality be sufficient over the entire night for reliable arousal and respiratory event detection (Table 3).

**Table 2.**
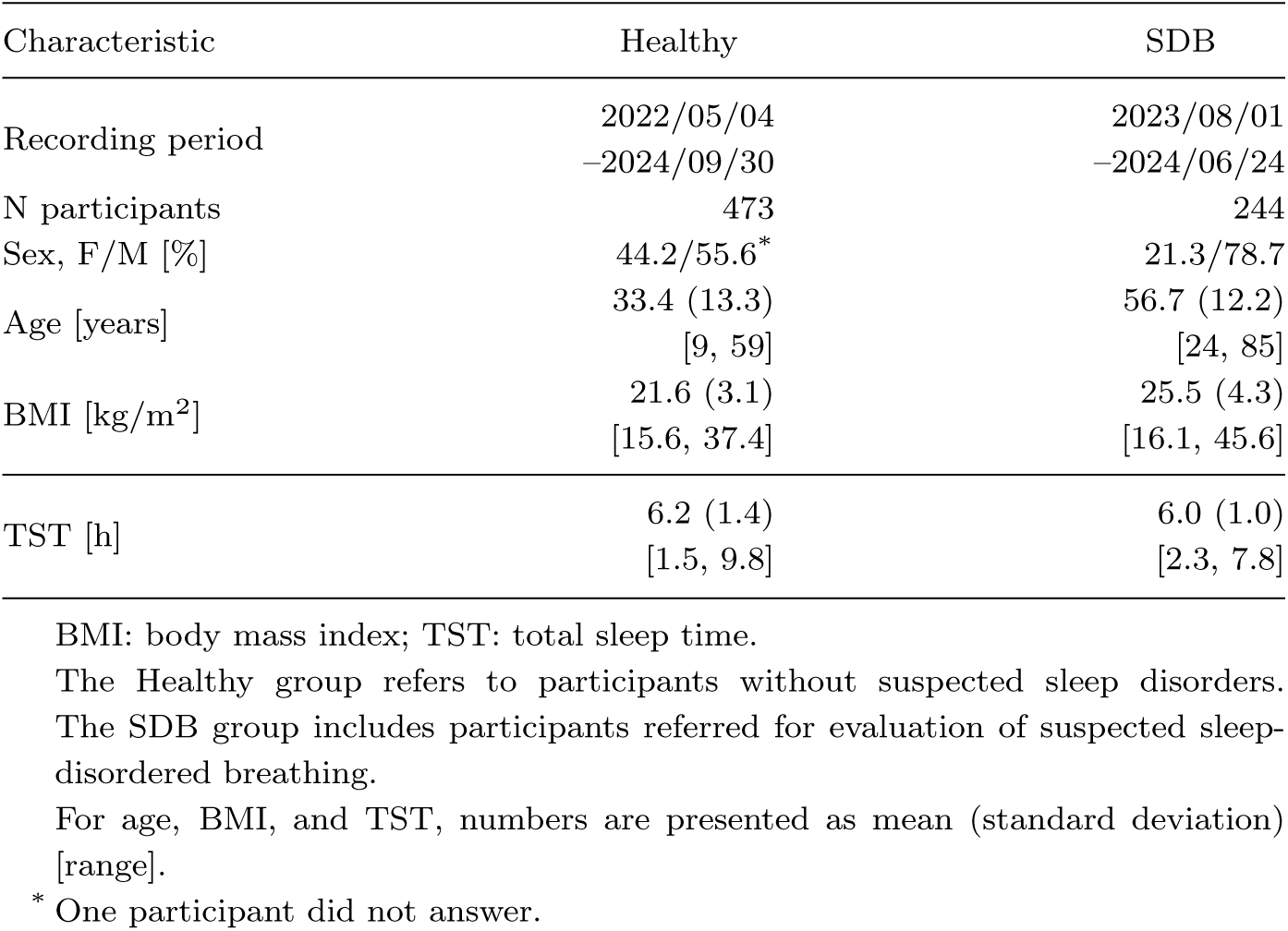
PSG datasets for the sleep stage study.

| Characteristic | Healthy | SDB |
| --- | --- | --- |
| Recording period | 2022/05/04<br>–2024/09/30 | 2023/08/01<br>–2024/06/24 |
| N participants | 473 | 244 |
| Sex, F/M [%] | 44.2/55.6* | 21.3/78.7 |
| Age [years] | 33.4 (13.3)<br>[9, 59] | 56.7 (12.2)<br>[24, 85] |
| BMI [kg/m<sup>2</sup>] | 21.6 (3.1)<br>[15.6, 37.4] | 25.5 (4.3)<br>[16.1, 45.6] |
| TST [h] | 6.2 (1.4)<br>[1.5, 9.8] | 6.0 (1.0)<br>[2.3, 7.8] |

BMI: body mass index; TST: total sleep time.
The Healthy group refers to participants without suspected sleep disorders. The SDB group includes participants referred for evaluation of suspected sleep-disordered breathing.
For age, BMI, and TST, numbers are presented as mean (standard deviation) [range].
\* One participant did not answer.

**Table 3.** PSG datasets for the arousal and respiratory event studies.

| Characteristic | Healthy | SDB |
| --- | --- | --- |
| Recording period | 2022/05/04<br>–2024/09/30 | 2023/08/01<br>–2024/06/24 |
| N participants | 412 | 178 |
| Sex, F/M [%] | 44.4/55.6 | 19.7/80.3 |
| Age [years] | 33.5 (13.2)<br>[9, 59] | 56.5 (12.4)<br>[24, 85] |
| BMI [kg/m<sup>2</sup>] | 21.6 (3.1)<br>[15.6, 32.3] | 25.2 (4.3)<br>[16.1, 45.6] |
| TST [h] | 6.2 (1.4)<br>[1.5, 9.8] | 6.0 (1.0)<br>[2.3, 7.8] |
| ArI [1/h] | 14.8 (9.1)<br>[2.1, 72.5] | 41.1 (17.6)<br>[6.2, 114.5] |
| AHI [1/h] | - | 40.9 (20.6)<br>[3.5, 103.0] |

BMI: body mass index; TST: total sleep time; ArI: arousal index; AHI: apnea–hypopnea index.
The Healthy group refers to participants without suspected sleep disorders. The SDB group includes participants referred for evaluation of suspected sleep-disordered breathing.
For age, BMI, TST, ArI, and AHI, numbers are presented as mean (standard deviation) [range].

For the respiratory event detection study, only recordings from participants with suspected SDB were used.

For the evaluation of inter-scorer agreement, 27 healthy recordings were used in the consensus study for sleep stage classification and arousal detection, and 5 SDB recordings were used in the respiratory event consensus study (Table 4).

**Table 4.** PSG datasets for the consensus studies.

| Characteristic | Healthy | SDB |
| --- | --- | --- |
| Recording period | 2022/05/04<br>–2022/10/29 | 2023/08/01<br>–2023/08/16 |
| N participants | 27 | 5 |
| Sex, F/M [%] | 29.6/70.4 | 0.0/100.0 |
| Age [years] | 40.1 (11.5)<br>[23, 59] | 56.8 (13.6)<br>[39, 73] |
| BMI [kg/m<sup>2</sup>] | 22.1 (2.0)<br>[18.8, 26.0] | 24.9 (4.7)<br>[20.3, 32.5] |
| TST [h] | 6.7 (1.2)<br>[4.8, 9.5] | 5.5 (0.5)<br>[5.1, 6.1] |
| ArI [1/h] | 13.3 (5.3)<br>[2.6, 26.2] | 19.7 (7.6)<br>[12.9, 29.4] |
| AHI [1/h] | - | 32.1 (16.7)<br>[13.4, 51.2] |

BMI: body mass index; TST: total sleep time; ArI: arousal index; AHI: apnea–hypopnea index.
The Healthy group refers to participants without suspected sleep disorders. The SDB group includes participants referred for evaluation of suspected sleep-disordered breathing.
For age, BMI, TST, ArI, and AHI, numbers are presented as mean (standard deviation) [range].

Table 5 summarizes the PSG recordings used for scorer calibration. For sleep stage and arousal scoring, calibration was conducted using 30 recordings from healthy participants and 7 recordings from the Calibration SDB group. Of the 30 healthy recordings, 25 were included in the consensus subset used in this study. The remaining five were not included in model development or evaluation: three were excluded because they were duplicate recordings from participants already represented elsewhere in the dataset, and two were excluded because part of the recording contained noise that made scoring impossible in some intervals. In addition, two further healthy recordings were later scored by all four scorers for monitoring purposes and were included in the consensus subset.

**Table 5.**
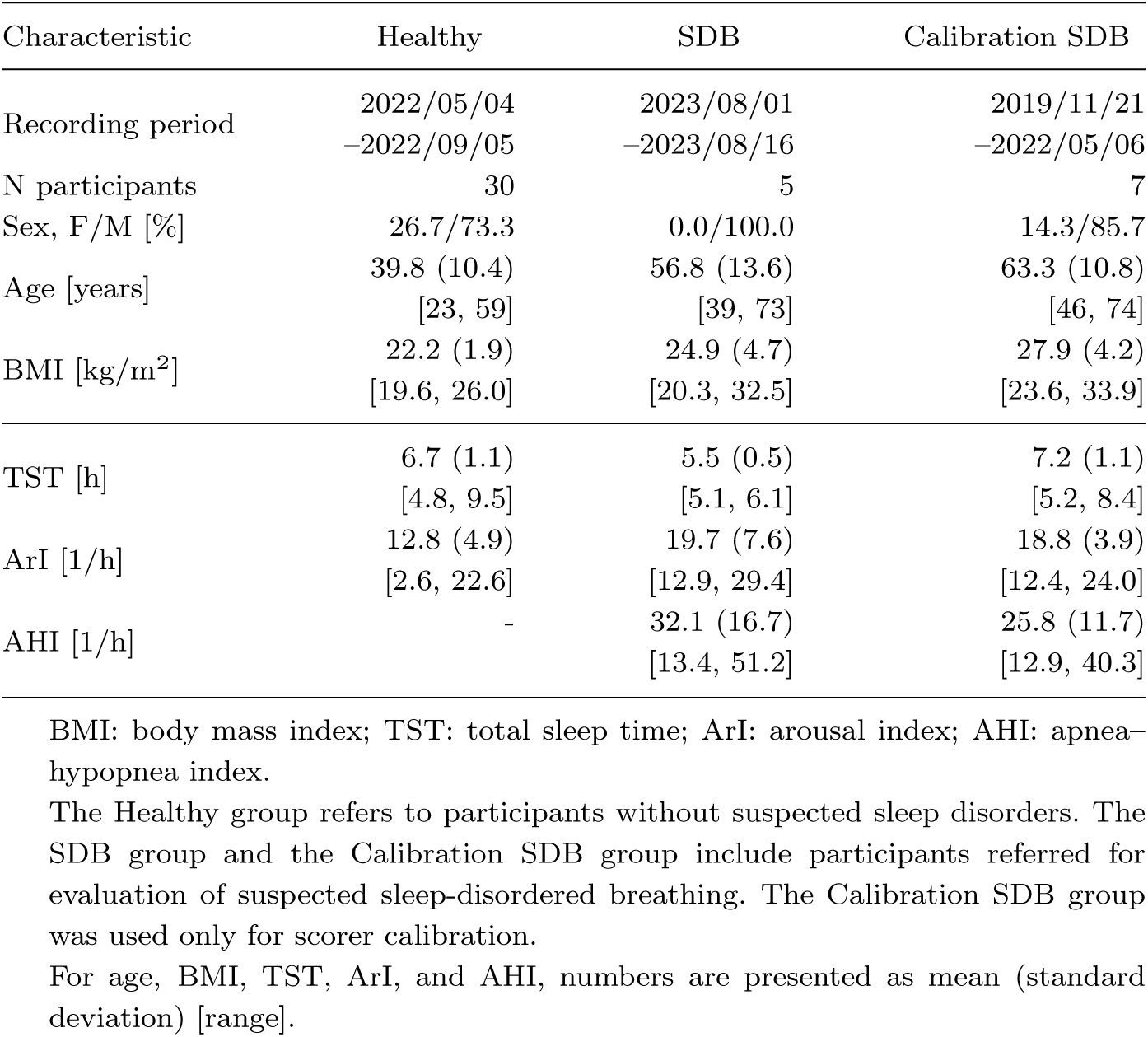
PSG datasets used for scorer calibration.

For respiratory event calibration, 5 recordings from the SDB group and 7 recordings from the Calibration SDB group were used. The 5 SDB recordings were the same subset as those used in the respiratory event consensus study.

### Annotation

Annotation of sleep stages, arousals, and respiratory events followed the AASM Scoring Manual. While the current manual is Version 3 [9], scoring in this study was conducted according to Version 2.6 [50], which was the version routinely used in Japan at the time. For each recording, the lights out and lights on times were first identified, and scoring was performed only within the interval between lights out and lights on.

Annotation was performed by four experienced scorers, each affiliated with a different institution. For sleep stage scoring, scorer calibration was conducted using recordings from the Healthy group together with recordings from the Calibration SDB group, as summarized in Table 5. The target was to achieve at least 85% agreement with the lead scorer for each sleep stage in each recording, while for recordings from the Calibration SDB group, the minimum target agreement was set at 80%. Each scorer first independently scored the calibration recordings. Segments with substantial disagreement were then reviewed with the lead scorer, and recordings with frequent disagreements were selectively rescored, with up to approximately four rounds of rescoring performed when needed.

For arousal scoring, scorer calibration was conducted using the same calibration recordings, and the resulting agreement was considered sufficient for the purposes of this study.

For respiratory event scoring, calibration was performed with a target event-level agreement of approximately 80% with the lead scorer. After an initial round of scoring, discrepant events were reviewed with the lead scorer, and depending on the recording, rescoring was repeated approximately three or four times.

Ater calibration, the remaining PSG recordings were distributed among the scorers, and each scorer independently annotated the assigned recordings. Two additional healthy recordings were also scored by all four scorers for sleep stage and arousal scoring and were included in the consensus dataset.

Except for the consensus subset, all respiratory event annotations were performed by a single expert scorer. Respiratory events included hypopnea, obstructive apnea, central apnea, and mixed apnea; however, for the purposes of this study, all were treated collectively as a single class of respiratory events.

Because PSG data were collected over an extended period, annotation sessions were also conducted at different times, which may have introduced minor temporal variations in scoring style.

### Algorithms

This study employed a GBDT-based framework for three tasks: sleep stage classification, arousal detection, and respiratory event detection. All tasks were formulated as epoch-wise classification problems based on manually annotated PSG signals. Data were segmented into fixed length epochs: 30-seconds epochs for sleep stage classification and 1-second epochs for arousal and respiratory events. From each epoch, task specific sets of time domain and frequency domain features were extracted from EEG, EOG, EMG, nasal pressure airflow, and SpO_2_ signals. Although the PSG recordings were acquired at different native sampling frequencies (Table 1), the feature extraction procedures operated on time-normalized segments and predefined frequency bands, ensuring that signals from both devices yielded harmonized feature representations.

A hierarchical model structure was used: the estimated sleep stage from the sleep stage model was included as a feature for arousal detection, and both the estimated sleep stage and arousal labels were used as features for respiratory event detection. To capture temporal dependencies in physiological signals, features from the target epoch and neighboring epochs were concatenated to form an extended context window.

All models were implemented using LightGBM [51], chosen for their suitability for heterogeneous tabular features and efficient training. For arousal and respiratory events, the 1-second epoch-wise probabilities output by the classifier were merged using task specific post-processing procedures to reconstruct contiguous events.

The following subsections describe the task specific feature sets, temporal context structures, classification models, and post-processing steps. Throughout all tasks, airflow features refer specifically to nasal pressure airflow derived from a cannula sensor, which was the airflow modality used in this study.

### Sleep Stage Classification

#### Sleep Stage Feature Extraction

Data were segmented into consecutive, non-overlapping 30-second epochs. For each epoch, a total of 1450 features were computed from EEG, EOG, and EMG channels.

For the EEG signals (six channels), power spectral density (PSD) features were computed using the Welch method [52], which estimates the power spectrum by averaging modified periodograms of overlapping signal segments to reduce variance. Each 30-second epoch was divided into six 5 second subsegments, and the log power spectral density was estimated for the following frequency bands: slow delta (0.5–2 Hz), fast delta (2–4 Hz), theta (4–8 Hz), alpha (8–13 Hz), beta (13–30 Hz), and spindle (11–16 Hz). Band power ratios were also derived. Identical PSD and ratio features were computed from the PSD estimated over the full 30-second epoch. Time domain features included the mean, root mean square (RMS), skewness, kurtosis, and 0.75 quantile for each channel.

For the EOG signals (two channels), PSD features were extracted at 1 Hz resolution from 0–35 Hz using the same six segment structure as for EEG. Time domain statistics (mean, RMS, skewness, kurtosis, 0.75 quantile) were also computed. For the EMG signal (one channel), RMS values were computed for each 1-second segment within the epoch, and the sequence of RMS values served as EMG features.

#### Sleep Stage Classification Model and Temporal Context

To incorporate temporal information, features from the target epoch and the two preceding and two following epochs were concatenated, forming a five-epoch context window (7250 features per input sample). LightGBM was used to classify each epoch into Wake, N1, N2, N3, or REM.

### Arousal Detection

#### Arousal Feature Extraction

For arousal detection, data were segmented into 1-second epochs, and labels were generated by converting event-based arousal annotations into 1-second epoch-wise labels.

For each epoch, 34 features were computed from EEG, EOG, EMG, nasal pressure airflow, and SpO_2_ signals, in addition to the estimated sleep stage.

The sleep stage estimated by the sleep stage model was included as a categorical variable and converted into dummy variables for N1, N2, N3, and REM.

For the EEG signals, PSD features were computed from each 1-second segment using the same spectral bands defined for sleep stage classification. From these, only beta power, spindle power, and the theta/alpha ratio were retained, based on feature importance analysis indicating their relevance for arousal discrimination. RMS was also computed for each EEG channel.

For the EOG and EMG channels, the RMS of each 1-second segment was extracted.

For the nasal pressure airflow signal, respiratory volume per time (RVT) was computed using a Hilbert-based method that estimates instantaneous respiratory amplitude and period [53] and the RVT values were then averaged within each 1- second segment.

For the SpO_2_ signal, the RMS and mean of each 1-second segment were used.

#### Arousal Classification Model and Temporal Context

Features from the target epoch and the 30 preceding and 30 following epochs were concatenated to form a 61 epoch context window (2074 features per sample). LightGBM was used to classify each epoch as arousal or non-arousal.

#### Arousal Post-Processing

Epochs with predicted arousal probability above 0.3 were marked as arousal candidates. Gaps of 5 seconds or less between candidates were bridged to form continuous events. Events shorter than 3 seconds were removed. Thresholds were selected by minimizing the mean absolute error between predicted and annotated Arousal Index (ArI).

### Respiratory Event Detection

#### Respiratory Event Feature Extraction

For respiratory event detection, data were segmented into 1-second epochs, and respiratory annotations were converted into epoch-wise labels following the same procedure as for arousals. For each epoch, seven features were computed from the estimated sleep stage, estimated arousal labels, nasal pressure airflow, and SpO_2_ signals.

The estimated sleep stage was included as dummy variables (N1, N2, N3, REM). The arousal detection model provided a binary arousal indicator (1 for predicted arousal, 0 otherwise).

For the nasal pressure airflow signal, RVT was computed for each 1-second segment as described in the Arousal Feature Extraction section. For the SpO_2_ signal, the mean of each 1-second segment was included.

#### Respiratory Event Classification Model and Temporal Context

A temporal context window of 61 seconds was constructed by concatenating features from the target epoch with the 30 preceding and 30 following epochs (427 features per input sample). LightGBM was used to classify each epoch as either a respiratory event or a non-event.

#### Respiratory Event Post-Processing

Epochs with predicted probability above 0.7 were marked as respiratory event candidates. Gaps of 2 seconds or less between candidate epochs were bridged to form continuous events. Events shorter than 10 seconds were removed. Thresholds were optimized by minimizing the mean absolute error in apnea–hypopnea index (AHI) compared with manual annotations.

### Model Training and Evaluation

#### Cross-Validation

All models were trained and evaluated using a participant level five-fold cross-validation (CV) framework. Because the dataset comprised healthy individuals and participants with suspected SDB, the two groups were handled separately when constructing the folds. Within each group, participants were randomly divided into five approximately equal subsets. Five final folds were then created by pairing the corresponding healthy and SDB subsets, ensuring comparable proportions of healthy and SDB recordings in each fold.

In each CV iteration, three folds were used for training, one fold served as the validation set for early stopping, and the remaining fold was held out for testing. For arousal detection and respiratory event detection, the same participant level fold assignments established for sleep stage classification were retained. Each participant thus had a consistent fold index (folds 1–5) across all tasks, while the actual recordings used for arousal or respiratory event detection corresponded to the task specific subsets. This ensured consistent participant grouping across tasks and prevented information leakage when sleep stage or arousal predictions were used as input features in downstream models.

Unless otherwise specified, all performance metrics reported in the Results section were computed on the held out test fold in each CV iteration and averaged across the five folds.

#### Sleep Stage Evaluation

To evaluate sleep stage classification performance, true and predicted sleep stages were compared for all 30-second epochs. From these epoch-wise labels, metrics (accuracy, Cohen’s kappa coefficient, macro averaged F_1_-score, and stage-wise F_1_-score for Wake, N1, N2, N3, and REM) were computed for each test fold and then averaged across the five folds.

At the recording level, total sleep time (TST) was calculated separately from the reference annotations and from the model outputs by summing the durations of all epochs labeled as N1, N2, N3, or REM for each recording. For each recording, the residual was defined as predicted TST minus true TST. Agreement between true and predicted TST was then summarized using residual plots and Bland–Altman limits of agreement (LoA) [54].

#### Event-Based Metrics

For arousal and respiratory event detection, event-based performance was evaluated by treating each true event (reference annotation) and each predicted event (model output) as a time interval defined by its onset and duration. For every pair of true and predicted events, the intersection over union (IoU) metric was computed as the ratio of the duration of their temporal overlap to the duration of their union [55]. A predicted event was considered a match if its IoU with a true event was at least 0.3. Matched pairs were counted as true positives (TP), true events without a matching prediction (IoU *≤* 0.3) were counted as false negatives (FN), and predicted events without a corresponding true event were counted as false positives (FP). The IoU threshold of 0.3 was chosen to align with previously proposed evaluation protocols for respiratory event detection, allowing direct comparison with prior human AI benchmarking studies [45].

From TP, FP, and FN, precision, recall, and F_1_-score were calculated for each test fold and then averaged across the five folds.

At the recording level, ArI and AHI were computed from both reference annotations and model outputs by dividing the total number of detected events by the TST in hours. For each recording, the residual was defined as predicted index minus true index.

#### Consensus Annotations and Human-Model Comparison

Consensus subsets were defined a priori for inter-scorer and human model comparison analyses. For sleep stage classification and arousal detection, 27 PSG recordings from healthy participants were annotated independently by all four scorers (Table 4). For respiratory event detection, five recordings from participants with suspected SDB were annotated by all four scorers.

For sleep stage classification, each scorer’s annotations were compared against consensus annotations derived from other scorers’ labels [56]. The model outputs were compared against consensus annotations created from the three highest accuracy scorers out of four [44].

For arousal and respiratory event detection, event-based annotations on the consensus subset were converted to 1-second epoch-wise binary labels (arousal vs. non-arousal) for each scorer. For each epoch, a majority vote among the three selected scorers (at least two arousal labels) was required to label the epoch as arousal, and consecutive arousal labeled epochs were merged into continuous events, yielding consensus arousal annotations [36]. Each scorer’s annotations were compared against consensus annotations derived from the other scorers’ labels. The model outputs were compared against consensus annotations created from the three highest accuracy scorers out of four.

For the consensus analyses, all metrics were computed based on each recording in the consensus subset and then averaged across recordings.

## Results

### Performance Metrics

#### Sleep Stage Classification

Table 6 summarizes the normalized confusion matrix across all folds.

**Table 6.** Normalized confusion matrix for sleep stage classification.

| True \ Predicted | Wake | N1 | N2 | N3 | REM |
| --- | --- | --- | --- | --- | --- |
| Wake | 89.3 | 7.6 | 0.9 | 0.1 | 2.0 |
| N1 | 6.3 | 68.6 | 19.6 | 0.1 | 5.4 |
| N2 | 0.4 | 6.4 | 86.9 | 4.7 | 1.5 |
| N3 | 0.1 | 0.1 | 14.4 | 85.4 | 0.0 |
| REM | 1.6 | 6.1 | 3.6 | 0.1 | 88.7 |

Values are expressed as row-wise percentages.
True labels (reference annotations) are shown in rows, and predicted labels (model outputs) are shown in columns.

Across the test folds, the model achieved the following performance, reported as mean (standard deviation; SD): accuracy of 0.840 (0.006), Cohen’s kappa of 0.791 (0.007), and an overall F_1_-score of 0.841 (0.005). Stage-wise F_1_-scores for Wake, N1, N2, N3, and REM were 0.890 (0.012), 0.711 (0.004), 0.848 (0.005), 0.865 (0.009), and 0.892 (0.006), respectively. These results indicate strong discrimination for Wake, N2, N3, and REM, whereas the lower performance observed for N1 is consistent with previous PSG-based classification studies.

Figure 1 shows the residual distribution for TST estimates. TSTs were calculated for each recording by summing the durations of N1, N2, N3, and REM stages. Residuals were symmetrically distributed around zero without a systematic bias, and the LoA were approximately *±*0.5 hours.

**Fig. 1:**
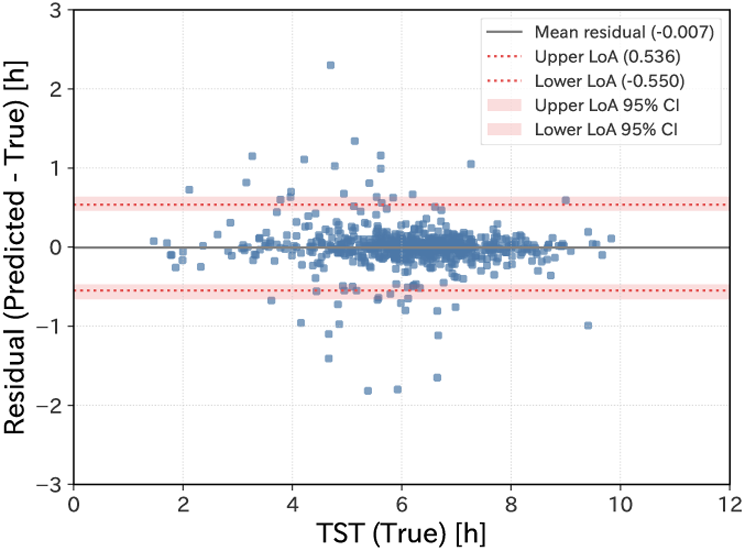
Residual plot for total sleep time (TST). The horizontal axis represents the true TST (from reference annotations) in hours, and the vertical axis represents the residual, defined as predicted TST (from model output) minus true TST. Each point corresponds to an individual recording. The solid horizontal line indicates the mean residual. The upper and lower dashed lines represent the limits of agreement (LoA), calculated as the mean residual *±* 1.96 SD of the residuals. The shaded regions indicate the 95% confidence intervals for the mean and LoA lines.

### Arousal Detection

Event-based performance was evaluated using the IoU-based matching criterion defined in the Model Training and Evaluation section. The model achieved mean (SD) recall, precision, and F_1_-scores of 0.725 (0.010), 0.742 (0.018), and 0.733 (0.011), respectively. These results indicate balanced sensitivity and specificity, with stable performance across folds.

Figure 2 shows the residual plot for the ArI. The LoA were approximately *±*15 events/h, and only a small number of outliers were observed. A slight tendency toward more negative residuals at higher ArI values may be present, although the number of observations in this range was limited.

**Fig. 2:**
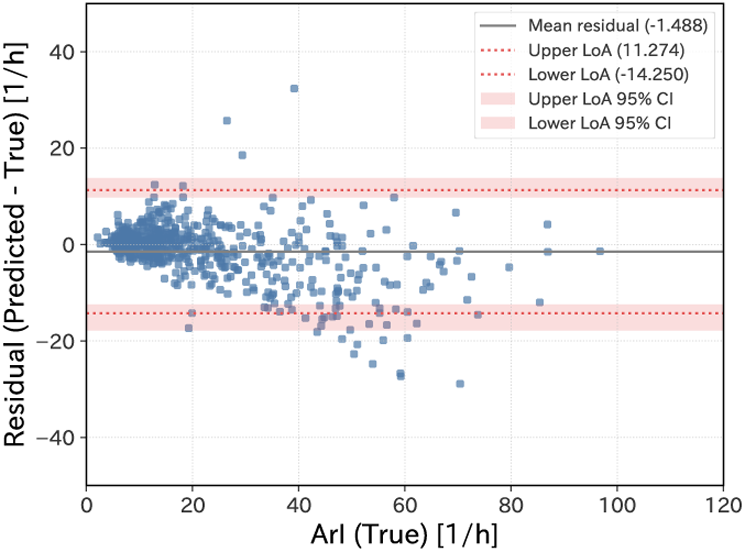
Residual plot for the arousal index (ArI). The horizontal axis represents the true ArI (from reference annotations), and the vertical axis represents the residual, defined as predicted ArI (from model output) minus true ArI. Each point corresponds to an individual recording. The solid horizontal line indicates the mean residual. The upper and lower dashed lines represent the limits of agreement (LoA), calculated as the mean residual *±* 1.96 SD of the residuals. The shaded regions indicate the 95% confidence intervals for the mean and LoA lines.

### Respiratory Event Detection

Respiratory event detection employed the same IoU-based matching and performance metrics as arousal detection. The model achieved mean (SD) recall, precision, and F_1_-scores of 0.829 (0.010), 0.807 (0.016), and 0.818 (0.012), respectively.

Figure 3 shows the residual plot for the AHI. Residuals were centered around zero with no systematic bias across the range of AHI values. LoAs were approximately *±*15 events/h, and only a few outliers were observed in the high AHI range.

**Fig. 3:**
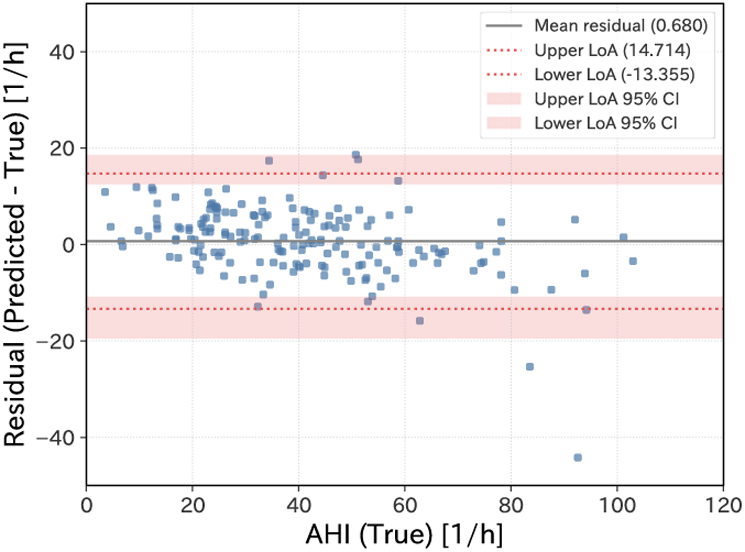
Residual plot for the apnea–hypopnea index (AHI). The horizontal axis represents the true AHI (from reference annotations), and the vertical axis represents the residual, defined as predicted AHI (from model output) minus true AHI. Each point corresponds to an individual recording. The solid horizontal line indicates the mean residual. The upper and lower dashed lines represent the limits of agreement (LoA), calculated as the mean residual *±* 1.96 SD of the residuals. The shaded regions indicate the 95% confidence intervals for the mean and LoA lines.

#### Consensus Study

##### Sleep Stage Classification Consensus

Consensus analysis for the sleep stage classification task was performed on the 27 recording subset described in the Consensus Annotations and Human-Model Comparison section. Metrics were obtained for each recording, and means and SD were calculated across the recordings. Tables 7 and 8 summarize the overall and stage wise performance metrics for individual scorers and the model against the consensus labels. Figure 4 shows the distribution of stage-wise F_1_-score across recordings.

**Fig. 4:**
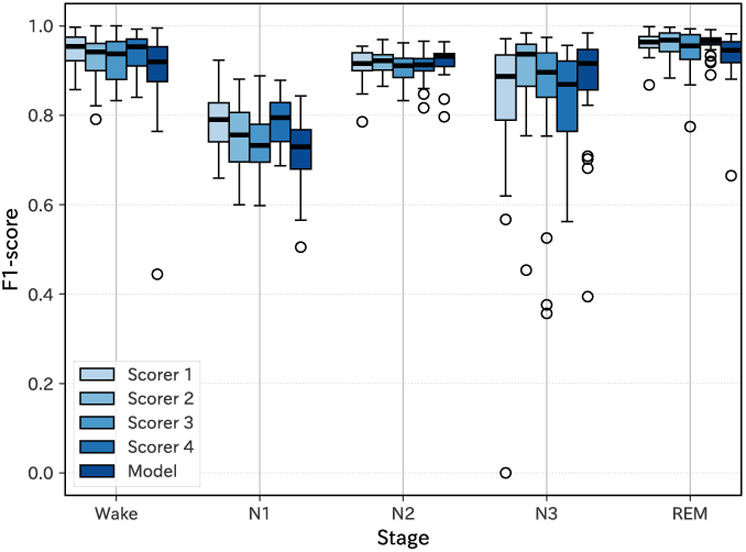
F_1_-score for each sleep stage of individual scorers and the model against consensus annotations. Boxplots showing the distribution of F_1_-score across the 27 recordings for each sleep stage (Wake, N1, N2, N3, REM) are presented for each scorer and the model. For each group, the box represents the interquartile range (IQR), with the thick horizontal line indicating the median. Whiskers extend to 1.5 *×* IQR or to the most extreme data points within that range, and individual points beyond the whiskers are plotted as outliers.

**Table 7.**
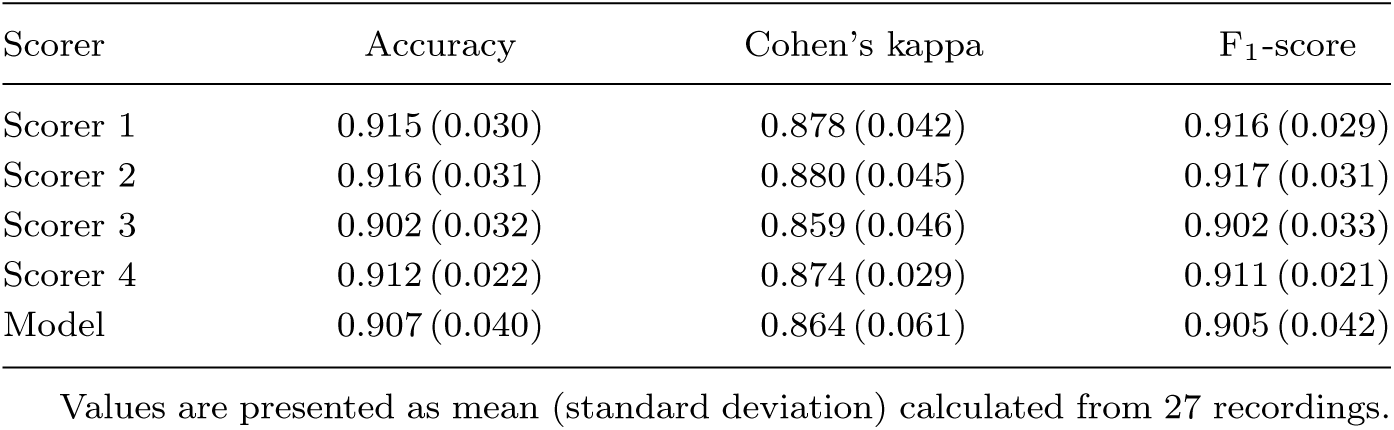
Performance metrics for sleep stage classification of individual scorers and the model.

**Table 8.** Stage wise F_1_-scores for sleep stage classification of individual scorers and the model.

| Scorer | Wake | N1 | N2 | N3 | REM |
| --- | --- | --- | --- | --- | --- |
| Scorer 1 | 0.947 (0.037) | 0.789 (0.061) | 0.912 (0.038) | 0.817 (0.199) | 0.961 (0.026) |
| Scorer 2 | 0.928 (0.053) | 0.751 (0.075) | 0.921 (0.027) | 0.890 (0.111) | 0.960 (0.030) |
| Scorer 3 | 0.925 (0.052) | 0.736 (0.069) | 0.905 (0.036) | 0.842 (0.167) | 0.946 (0.048) |
| Scorer 4 | 0.942 (0.040) | 0.783 (0.055) | 0.911 (0.034) | 0.838 (0.104) | 0.961 (0.024) |
| Model | 0.897 (0.103) | 0.713 (0.078) | 0.920 (0.035) | 0.874 (0.127) | 0.931 (0.060) |

Values are presented as mean (standard deviation) calculated from 27 recordings.

The model demonstrated comparable performance to that of human scorers across all sleep stages. The score for the N1 stage was lower than for other stages but still within the range of human scorer performance.

##### Arousal Detection Consensus

Consensus analysis for the arousal detection task was performed on the 27 recording subset described in the Consensus Annotations and Human-Model Comparison section. Event-based performance of each scorer and the model was evaluated against the consensus arousal annotations using the IoU-based matching criterion and event level metrics defined in the Model Training and Evaluation section. Table 9 summarizes recall, precision, and F_1_-score for individual scorers and the model, and Figure 5 shows recall and precision for each against the consensus annotations.

**Fig. 5:**
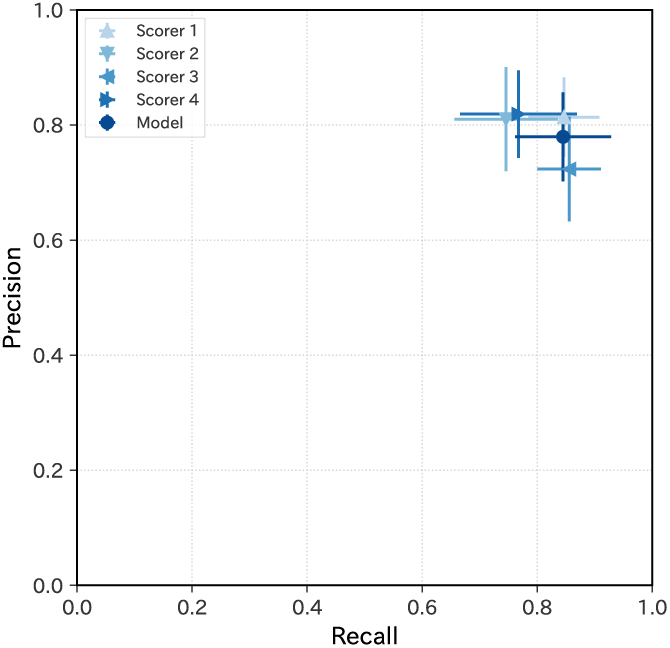
Recall and precision for arousal detection by individual scorers and by the model, evaluated against consensus annotations. Mean recall and precision across recordings are shown, with error bars indicating the corresponding standard deviations (SD).

**Table 9.** Performance metrics for arousal detection of individual scorers and the model.

| Scorer | Recall | Precision | $F_1$-score |
| --- | --- | --- | --- |
| Scorer 1 | 0.846 (0.062) | 0.814 (0.069) | 0.827 (0.047) |
| Scorer 2 | 0.746 (0.090) | 0.810 (0.090) | 0.770 (0.062) |
| Scorer 3 | 0.856 (0.055) | 0.724 (0.091) | 0.780 (0.061) |
| Scorer 4 | 0.768 (0.101) | 0.819 (0.076) | 0.786 (0.067) |
| Model | 0.845 (0.083) | 0.780 (0.078) | 0.807 (0.056) |

Values are presented as mean (standard deviation) calculated from 27 recordings.

The model demonstrated comparable performance to that of human scorers.

##### Respiratory Event Detection Consensus

For respiratory event detection, consensus analysis was conducted on five recordings from participants with suspected SDB, as defined in the Consensus Annotations and Human-Model Comparison section. Consensus respiratory event annotations were compared with each scorer’s annotations and with the model outputs using the same IoU-based matching criterion and event-based metrics as for arousals. Table 10 summarizes recall, precision, and F_1_-score for individual scorers and the model, and Figure 6 illustrates recall and precision for each against the consensus annotations.

**Fig. 6:**
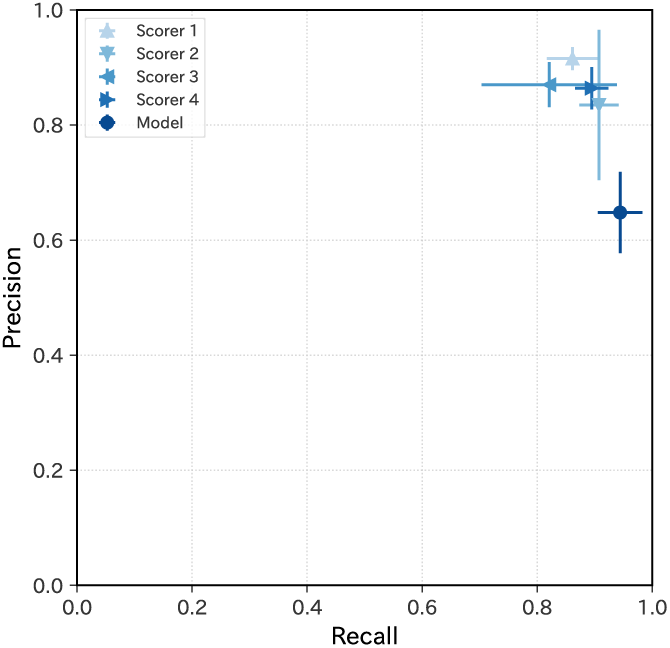
Recall and precision for respiratory event detection by individual scorers and by the model, evaluated against consensus annotations. Mean recall and precision across recordings are shown, with error bars indicating the corresponding standard deviations (SD).

**Table 10.** Performance metrics for respiratory event detection of individual scorers and the model.

| Scorer | Recall | Precision | F<sub>1</sub>-score |
| --- | --- | --- | --- |
| Scorer 1 | 0.861 (0.045) | 0.916 (0.020) | 0.888 (0.033) |
| Scorer 2 | 0.907 (0.034) | 0.835 (0.131) | 0.865 (0.078) |
| Scorer 3 | 0.821 (0.118) | 0.870 (0.039) | 0.842 (0.073) |
| Scorer 4 | 0.895 (0.029) | 0.864 (0.037) | 0.878 (0.011) |
| Model | 0.944 (0.039) | 0.648 (0.071) | 0.767 (0.054) |

Values are presented as mean (standard deviation) calculated from 5 recordings.

The model demonstrated slightly lower overall performance than human scorers, characterized by higher recall and lower precision in the consensus subset.

This recall–precision imbalance contrasts with the more balanced performance observed in the Respiratory Event Detection section, where precision and recall were closely aligned. The deviation was observed specifically in the consensus subset. The consensus subset consists of five recordings, and its performance metrics therefore reflect a limited evaluation sample. In addition, these recordings were annotated at an early stage of the study, whereas most annotations were generated later under the same calibration framework. These factors may be associated with the recall–precision imbalance observed in this subset.

## Discussion

### Overview of Model Performances

This study evaluated three models designed for distinct sleep related tasks: sleep stage classification, arousal detection, and respiratory event detection from PSG data. All models were implemented using GBDT architectures rather than deep learning frameworks and were designed to operate sequentially within a unified pipeline. Specifically, the arousal detection model leveraged outputs from the sleep stage classifier, and the respiratory event detection model further incorporated both sleep stage and arousal information. This hierarchical design reflects the dependency structure of physiological events during sleep and enables each downstream task to benefit from contextual information provided by preceding stages.

Detailed performance metrics for all tasks are summarized in the Performance Metrics section.

In sleep stage classification, prior work employing deep learning approaches has reported F_1_-scores of 0.90 (0.04), 0.53 (0.07), 0.85 (0.04), 0.76 (0.07), and 0.90 (0.02) for Wake, N1, N2, N3, and REM stages, respectively [44]. In the present study, comparable performance was achieved for the Wake, N2, and REM stages, while notably higher F_1_-scores were observed for the N1 (0.711 (0.004)) and N3 (0.865 (0.009)) stages. Notably, substantial variability in N1 performance across datasets (F_1_-scores ranging from 0.45 to 0.64) has been documented in prior work [44], underscoring the dataset dependent nature of this stage.

For arousal detection, previous studies have reported event level F_1_-scores ranging from 0.70 to 0.79 depending on the dataset [37]. The performance achieved in the present study (0.733 (0.011)) falls within this range.

With respect to respiratory event detection, earlier investigations of binary apnea–hypopnea detection have reported F_1_-scores between 0.517 and 0.665 [39]. In contrast, the present study achieved an F_1_-score of 0.818 (0.012). While differences in datasets and event definitions limit direct comparability, this result indicates that robust respiratory event detection is achievable even without deep-learning-based architectures.

Taken together, these findings extend earlier observations that non-deep-learning methods can perform competitively in sleep stage classification to event-based tasks [31]. By combining interpretable GBDT classifiers with task specific post-processing algorithms, the proposed models successfully estimated both the onset and duration of arousal and respiratory events with high temporal precision, while maintaining model interpretability and feasibility for real-world deployment. In addition, GBDT models offer advantages in terms of reproducibility, computational efficiency, and stable implementation in clinical and large-scale research environments [57, 58].

At the same time, direct quantitative comparisons across studies remain inherently challenging, as datasets, annotation protocols, and event definitions differ substantially. Accordingly, variability in reported performance across prior work highlights that achievable model accuracy is strongly influenced by dataset specific factors, particularly the characteristics and consistency of the underlying reference annotations. For this reason, the following section focuses on a detailed examination of annotation quality and consensus scoring in the present dataset, in order to more rigorously contextualize the observed model performance.

### Annotation Quality and Consensus Study

In this study, particular attention was paid to the quality and consistency of the reference annotations. Before scoring, all scorers participated in structured calibration sessions to align interpretation criteria and reduce subjective variability across scorers.

As described in the Consensus Study section, the consensus analyses showed that, although performance for respiratory event detection was slightly lower than human inter-scorer agreement, the proposed models achieved human level performance overall within the present dataset. This establishes an interpretable reference for model performance, as agreement among human experts effectively defines a practical performance ceiling under a given annotation protocol.

For sleep stage classification, a previous study using a deep-learning-based model reported F_1_-scores of 0.88 (0.10), 0.56 (0.14), 0.86 (0.05), 0.73 (0.23), and 0.93 (0.05) for Wake, N1, N2, N3, and REM, respectively, in healthy participants, indicating performance comparable to inter-scorer agreement [44]. In the present study, the proposed model achieved comparable overall performance, while exhibiting higher performance particularly in the N1 and N3 stages. These stages are widely recognized as difficult to score consistently among human experts and are known to exhibit substantial inter-scorer variability [12–16]. The resulting differences in stage-wise performance patterns suggest that variations in annotation consistency across datasets can substantially influence achievable model performance, especially for these challenging sleep stages.

For arousal and respiratory event detection, the consensus analysis was conducted using a protocol analogous to that employed in an earlier comparative study of human and automated scoring, with a primary focus on respiratory events [45]. That study reported a model F_1_-score of approximately 0.57 (0.23) for respiratory event detection, while human scorers achieved F_1_-scores in the range of 0.52–0.58. In contrast, in the present study, the model achieved an F_1_-score of 0.767 (0.054), and human scorers achieved F_1_-scores ranging from 0.842 to 0.888. This higher level of agreement among human scorers was accompanied by a corresponding shift in model performance, although a gap relative to human level performance remains. Arousal detection was not evaluated in the earlier comparative study; however, the present results similarly demonstrated comparable performance between the model and human scorers for arousal detection.

In the consensus-based comparison, respiratory event detection showed relatively high recall but lower precision, whereas the results based on the full dataset were more balanced. One possible explanation is that annotation practice may have shifted over time, such that the broader set of annotations used across model development and evaluation became relatively more permissive in identifying respiratory events than the earlier consensus subset. If so, the model would tend to detect respiratory events more readily, leading to increased false-positive detections when evaluated against the consensus reference. This pattern may reflect task-specific differences in annotation tendencies, but it does not change the broader interpretation of the results across tasks.

Across the evaluated tasks, these findings indicate that even relatively simple GBDT models can approach or achieve human level performance. At the same time, the results indicate that higher levels of annotation quality are associated with higher achievable model performance when models are trained and evaluated on such data. This further highlights the central role of annotation quality in determining the achievable performance of automated sleep analysis systems. Moreover, explicitly characterizing human level agreement provides an essential reference for interpreting model performance, ensuring that reported improvements are interpreted relative to the inherent variability of human scoring rather than based solely on absolute performance metrics.

### Limitations

This study has several limitations that should be considered.

One limitation is that the datasets used in this work excluded recordings with extensive signal loss, severe noise, or missing channels. While this selection ensured reliable model training and evaluation, real-world clinical PSG recordings frequently contain artifacts, partial signal loss, or reduced signal quality. In routine practice, diagnostic interpretation must therefore be performed under imperfect conditions that are not fully represented in the present datasets [59–61].

Another limitation is that differences in hardware and software among PSG systems may affect signal characteristics. Although the AASM Scoring Manual [9] provides baseline technical specifications for signal acquisition, variations in hardware configurations, sensor types, filtering pipelines, and sampling rates across PSG systems can introduce systematic differences in recorded signals [62]. In the present study, PSG data acquired at different institutions using distinct recording systems were jointly analyzed, and stable performance was observed. However, because one dataset consisted exclusively of healthy participants while the other included participants with suspected sleep-disordered breathing, differences in participant characteristics were confounded with differences in recording systems and clinical settings. As a result, the isolated effects of hardware- or facility-related factors cannot be disentangled from population-related variability. In addition, some PSG systems have only three EEG channels. To generalize the present findings to such systems, future work may be extended to include datasets with limited EEG channels.

A further limitation concerns the annotations themselves. Although high-quality annotations were obtained through calibration sessions among four expert scorers, residual ambiguity in PSG scoring remains unavoidable. Previous studies have shown that even experienced scorers exhibit substantial inter-scorer variability and that calibration can improve agreement without fully eliminating disagreement [63]. Moreover, ambiguities in the AASM scoring rules allow room for differing interpretations. In addition, manual scoring in this study was based on Version 2.6 of the AASM Scoring Manual [50], whereas the current version is Version 3 [9]. Because Version 3 includes updates to some scoring rules, direct comparison with studies scored strictly according to the current manual should be made with caution. However, this difference is not considered to alter the main interpretation of the present study, which focuses on the effect of annotation quality on model evaluation. Inter-scorer agreement has been reported to be higher among scorers from the same clinical centers or countries than across different institutions or regions [64–66], suggesting the influence of shared local scoring conventions and institutional practices.

The calibration procedure may also have influenced the inter-scorer agreement estimates, because some of the same recordings were used both for calibration and for the consensus study, and epochs or events reviewed during calibration may have been scored more consistently thereafter. However, only a limited portion of each recording was subjected to detailed review, and most epochs or events were not directly affected by this process.

Related to the annotation quality, recent evaluations have reported that the performance of some automatic scoring systems remains insufficient for routine clinical use [67]. Such performance degradation may partly reflect mismatches in scoring conventions between training and evaluation datasets, rather than limitations of the underlying algorithms [68]. Accordingly, the present model may not achieve the same level of agreement when applied to independently annotated datasets, even when those datasets exhibit high internal inter-scorer reliability. This underscores the importance of external validation using datasets collected at different institutions and annotated by distinct scorer groups.

The study population consisted exclusively of Japanese participants. Physiological and demographic differences may influence PSG signal characteristics. In particular, skin pigmentation has been shown to affect pulse oximetry measurements [69–71]. Therefore, the generalizability of the present findings to other populations cannot be assumed, and validation in more diverse cohorts is required.

Finally, the present framework still has important scope-related limitations for comprehensive PSG interpretation. The respiratory event detection model treated all respiratory events as a single category and did not distinguish between apnea and hypopnea or between obstructive and central events, although such subtyping is clinically important for diagnosis and treatment planning [72, 73]. In addition, respiratory event detection relied primarily on the nasal pressure airflow signal and did not incorporate thermistor airflow or thoracoabdominal effort signals as model inputs. Both nasal pressure and thermistor are used in PSG to assess airflow reduction; however, in the present dataset, nasal pressure was more consistently recorded with acceptable signal quality than thermistor, and the final model was therefore developed using nasal pressure as the primary airflow input. This was considered a practical configuration for the present task, which was limited to detecting the presence or absence of respiratory events. By contrast, thoracoabdominal effort signals are particularly important for differentiating obstructive from central respiratory events, but these signals were not included because such subtype classification was outside the scope of the current model. Nevertheless, reliance on nasal pressure alone may reduce robustness when the pressure signal is degraded and may limit generalizability to datasets acquired with different airflow sensor configurations. In addition, periodic limb movements (PLM) [74], which are also an important component of comprehensive PSG interpretation alongside sleep stages, arousals, and respiratory events, were not analyzed because PLM-related annotations were not available in the current dataset. More comprehensive future development should therefore include evaluation of the complementary use of thermistor and thoracoabdominal effort signals, expansion of the analyzed channel set where appropriate, and incorporation of PLM annotation and detection.

### Clinical Implications and Future Directions

For practical deployment, explicit handling of real-world signal artifacts is essential. Numerous studies have proposed methods for detecting artifacts in PSG signals, including approaches targeting EEG contamination, movement-related artifacts, and corruption of respiratory signals [75–78]. Such methods could be integrated into preprocessing pipelines prior to sleep staging or respiratory event detection. At the same time, there remain cases in which automated systems may struggle, whereas such cases can be readily recognized by experienced human scorers [79]. These findings further emphasize the necessity of incorporating expert review into clinical workflows. Several studies have demonstrated that combining automated algorithms with expert review can be effective in clinical PSG workflows [80–82]. Semi-automated approaches have been shown to reduce scoring time while maintaining or improving agreement with expert consensus by allowing clinicians to focus on difficult or ambiguous segments.

Beyond signal quality, variability in PSG interpretation also arises from inter-scorer disagreement [12–16] and from local scoring conventions that develop within specific institutions or regional contexts [64–66]. Such variability complicates structured discussion of discrepant cases, as human judgments may vary across scorers and evolve over time.

In this setting, stable and reproducible model outputs may provide a useful reference for identifying scorer-related ambiguity. By comparing human judgments against consistent model predictions, ambiguous cases can be highlighted more explicitly, potentially supporting clarification and calibration of scoring decisions [83]. At the same time, models trained on institution-specific data may inherit local biases. From a practical standpoint, retraining or fine-tuning models on local datasets may allow alignment with institution-specific scoring conventions when such adaptation is clinically desirable [84, 85].

With respect to dataset size, prior work has shown that the performance of automated sleep staging models tends to plateau once the number of training recordings exceeds several hundred, with additional data contributing primarily to increased heterogeneity rather than improved accuracy [86]. The present study includes a dataset size comparable to or larger than those reported to reach this performance plateau, suggesting that the available data volume is sufficient for evaluating the proposed approach.

Importantly, further increases in dataset size may not necessarily lead to performance gains if additional data introduce greater heterogeneity in patient populations, recording systems, or annotation practices. In particular, incorporating datasets from different institutions or regions may add variability arising from local scoring conventions and implicit annotation differences, potentially reducing overall agreement. Future data expansion should therefore emphasize careful curation, harmonization of annotation practices, and strategies that explicitly address domain and annotation heterogeneity, rather than indiscriminate aggregation of additional recordings.

In this context, approaches that explicitly characterize scoring uncertainty may further strengthen collaboration between human experts and automated systems. Methods that highlight epochs associated with intrinsically ambiguous sleep stage transitions or low model confidence can guide targeted expert review and calibration. Recent work on probabilistic and uncertainty-aware sleep staging frameworks suggests that such representations provide quantitative insight into inter-scorer disagreement and may support improved scoring consistency over time [46–48]. More broadly, recent perspectives on advanced PSG analysis have emphasized the potential of moving beyond conventional rule-based representations toward integrative frameworks that capture multidimensional sleep dynamics across time, signals, and physiological domains, thereby enabling analyses that are difficult to achieve through visual inspection alone [87].

At a system level, federated learning has emerged as a promising framework for privacy-preserving collaboration across institutions. Recent studies have demonstrated that federated approaches to sleep stage classification can achieve performance comparable to centralized training while allowing data to remain within local clinical environments [88–90]. Such strategies may facilitate large-scale validation and model refinement across institutions while mitigating data-sharing constraints.

Finally, despite these methodological advances, translating automated PSG analysis into routine clinical practice remains challenging. Prior work has highlighted practical barriers such as integration with existing clinical workflows, user trust and acceptance, interpretability of model outputs, and the need for robust validation under real-world conditions, all of which must be systematically addressed to enable safe and effective clinical adoption [91].

Taken together, the present results demonstrate that models trained on carefully curated and internally consistent PSG datasets can achieve high performance with relevance to clinical PSG analysis across multiple scoring tasks. At the same time, effective deployment in real-world clinical environments requires explicit consideration of signal artifacts, scorer-related variability, and local practice differences. Accordingly, automated PSG analysis is best implemented as part of an integrated system that supports expert review and decision-making, rather than as a standalone replacement, with continued emphasis on usability, transparency, and seamless integration into clinical workflows. In addition, practical adoption will depend on thoughtful system design that facilitates efficient interaction between automated outputs and human expertise, enabling clinicians to readily interpret, review, and refine model-assisted results in daily practice.

## Data Availability

The data that support the findings of this study are not publicly available due to ethical, legal, and contractual restrictions related to participant privacy and data ownership.

## Acknowledgements

The authors thank Ms. Sayaka Chino, Mr. Kazuyoshi Namba, and Ms. Masako Sato for their invaluable contributions to the annotation of the polysomnography recordings. The authors appreciate Mr. Noboru Oki of Y.K. Norupro Light Systems for his technical support during the data analysis process. The authors further acknowledge Dr. Koji Ode, Dr. Akifumi Kishi, and Dr. Hiroki Ueda of the University of Tokyo for their constructive discussions and perspectives that helped shape the conceptual foundation of this research.

Natural language processing tools were used to assist with language editing.

## Disclosure statement

Financial Disclosure: This research was designed, conducted, and funded by ACCELStars, Inc. All authors are financially affiliated with ACCELStars, Inc. in relation to this work.

Non-financial Disclosure: None.

Preprint repositories: This manuscript has also appeared as a preprint on medRxiv (https://doi.org/10.64898/2025.12.15.25342334).

## Data availability

The data underlying this article cannot be shared publicly due to ethical, legal, and contractual restrictions related to participant privacy and data ownership. The data will be shared on reasonable request to the corresponding author.

## Notes

### Funding Statement

This study did not receive any funding.

### Author Declarations

The Ethics Committee of Medical Corporation Kouseikai Sone Clinic gave ethical approval for this work (Protocol IDs: SUG08869 and SUG08884).

### Summary of Updates

Details of the calibration dataset were added.

## References

1. Fava C, Montagnana M, Favaloro E, Guidi G, Lippi G. Obstructive Sleep Apnea Syndrome and Cardiovascular Diseases. Seminars in Thrombosis and Hemostasis. 2011;37(03):280–97. 10.1055/s-0031-1273092.

2. Ong J, Crawford M. Insomnia and Obstructive Sleep Apnea. Sleep medicine clinics. 2013;8 3:389–98. 10.1016/j.jsmc.2013.04.004.

3. Juan ID, Carmenates LIP. Obstructive sleep apnea syndrome. Journal of Otolaryngology-ENT Research. 2023. 10.15406/joentr.2023.15.00533.

4. Lavalle S, Masiello E, Iannella G, et al. Unraveling the Complexities of Oxidative Stress and Inflammation Biomarkers in Obstructive Sleep Apnea Syndrome: A Comprehensive Review. Life. 2024;14. 10.3390/life14040425.

5. Benjafield AV, Ayas NT, Eastwood PR, Heinzer R, Ip MW, Morrell MJ. Estimation of the global prevalence and burden of obstructive sleep apnoea: a literature-based analysis. The Lancet Respiratory Medicine. 2019;7(8):687–98. 10.1016/S2213-2600(19)30198-5.

6. Chang JL, Goldberg AN, Alt JA, et al. International Consensus Statement on Obstructive Sleep Apnea. International Forum of Allergy & Rhinology. 2023;13(7):1061–482. 10.1002/alr.23079.

7. Kapur VK, Auckley DH, Chowdhuri S, et al. Clinical Practice Guideline for Diagnostic Testing for Adult Obstructive Sleep Apnea: An American Academy of Sleep Medicine Clinical Practice Guideline. Journal of Clinical Sleep Medicine. 2017;13(03):479–504. 10.5664/jcsm.6506.

8. Penzel T, Conradt R. Computer based sleep recording and analysis. Sleep Medicine Reviews. 2000;4(2):131–48. 10.1053/smrv.1999.0087.

9. American Academy of Sleep Medicine. The AASM Manual for the Scoring of Sleep and Associated Events: Rules, Terminology and Technical Specifications. Version 3 ed. Darien, IL: American Academy of Sleep Medicine;2023.

10. Caples SM, Anderson WM, Calero K, Howell M, Hashmi SD. Use of polysomnography and home sleep apnea tests for the longitudinal management of obstructive sleep apnea in adults: an American Academy of Sleep Medicine clinical guidance statement. Journal of Clinical Sleep Medicine. 2021;17(6):1287–93. 10.5664/jcsm.9240.

11. Berry RB, Abreu AR, Krishnan V, Quan SF, Strollo PJ, Malhotra RK. A transition to the American Academy of Sleep Medicine-recommended hypopnea definition in adults: initiatives of the Hypopnea Scoring Rule Task Force. Journal of Clinical Sleep Medicine. 2022;18(5):1419–25. 10.5664/jcsm.9952.

12. Danker-Hopfe H, Anderer P, Zeitlhofer J, et al. Interrater reliability for sleep scoring according to the Rechtschaffen & Kales and the new AASM standard. Journal of Sleep Research. 2009;18(1):74–84. 10.1111/j.1365-2869.2008.00700.x.

13. Rosenberg RS, Hout SV. The American Academy of Sleep Medicine Inter-scorer Reliability Program: Sleep Stage Scoring. Journal of Clinical Sleep Medicine. 2013;09(01):81–7. 10.5664/jcsm.2350.

14. Zhang X, Dong X, Kantelhardt JW, et al. Process and outcome for international reliability in sleep scoring. Sleep and Breathing. 2014;19(1):191–5. 10.1007/s11325-014-0990-0.

15. Cesari M, Stefani A, Penzel T, et al. Interrater sleep stage scoring reliability between manual scoring from two European sleep centers and automatic scoring performed by the artificial intelligence-based Stanford-STAGES algorithm. Journal of Clinical Sleep Medicine. 2021;17(6):1237–47. 10.5664/jcsm.9174.

16. Lee YJ, Lee JY, Cho JH, Choi JH. Interrater reliability of sleep stage scoring: a meta-analysis. Journal of Clinical Sleep Medicine. 2022;18(1):193–202. 10.5664/jcsm.9538.

17. Koley B, Dey D. An ensemble system for automatic sleep stage classification using single channel EEG signal. Computers in Biology and Medicine. 2012;42(12):1186–95. 10.1016/j.compbiomed.2012.09.012.

18. Khalighi S, Sousa T, Pires G, Nunes U. Automatic sleep staging: A computer assisted approach for optimal combination of features and polysomnographic channels. Expert Systems with Applications. 2013;40(17):7046–59. 10.1016/j.eswa.2013.06.023.

19. Zhang Y, Zhang X, Liu W, et al. Automatic Sleep Staging using Multi-dimensional Feature Extraction and Multi-kernel Fuzzy Support Vector Machine. Journal of Healthcare Engineering. 2014;5(4):410705. 10.1260/2040-2295.5.4.505.

20. Lajnef T, Chaibi S, Ruby P, et al. Learning machines and sleeping brains: Automatic sleep stage classification using decision-tree multi-class support vector machines. Journal of Neuroscience Methods. 2015;250:94–105. Cutting-edge EEG Methods. 10.1016/j.jneumeth.2015.01.022.

21. Giannakeas N. EEG-Based Automatic Sleep Stage Classification. Biomedical Journal of Scientific & Technical Research. 2018;7(4). 10.26717/bjstr.2018.07.001535.

22. Gunnarsdottir KM, Gamaldo CE, Salas RME, Ewen JB, Allen RP, Sarma SV. A Novel Sleep Stage Scoring System: Combining Expert-Based Rules with a Decision Tree Classifier. In: 2018 40th Annual International Conference of the IEEE Engineering in Medicine and Biology Society (EMBC);2018. p. 3240–3. 10.1109/EMBC.2018.8513039.

23. Supratak A, Dong H, Wu C, Guo Y. DeepSleepNet: A Model for Automatic Sleep Stage Scoring Based on Raw Single-Channel EEG. IEEE Transactions on Neural Systems and Rehabilitation Engineering. 2017;25(11):1998–2008. 10.1109/TNSRE.2017.2721116.

24. Perslev M, Jensen M, Darkner S, Jennum PJr, Igel C. U-Time: A Fully Convolutional Network for Time Series Segmentation Applied to Sleep Staging. In: Wallach H, Larochelle H, Beygelzimer A, d’Alché-Buc F, Fox E, Garnett R, editors. Advances in Neural Information Processing Systems 32. Curran Associates, Inc.;2019. p. 4415–26.

25. Eldele E, Chen Z, Liu C, et al. An Attention-Based Deep Learning Approach for Sleep Stage Classification With Single-Channel EEG. IEEE Transactions on Neural Systems and Rehabilitation Engineering. 2021;29:809–18. 10.1109/TNSRE.2021.3076234.

26. Li C, Qi Y, Ding X, Zhao J, Sang T, Lee M. A Deep Learning Method Approach for Sleep Stage Classification with EEG Spectrogram. International Journal of Environmental Research and Public Health. 2022;19(10):6322. 10.3390/ijerph19106322.

27. Phan H, Chén OY, Tran MC, Koch P, Mertins A, De Vos M. XSleepNet: Multi-View Sequential Model for Automatic Sleep Staging. IEEE Transactions on Pattern Analysis and Machine Intelligence. 2022;44(9):5903–15. 10.1109/TPAMI.2021.3070057.

28. Pradeepkumar J, Anandakumar M, Kugathasan V, et al. Toward Interpretable Sleep Stage Classification Using Cross-Modal Transformers. IEEE Transactions on Neural Systems and Rehabilitation Engineering. 2024;32:2893–904. 10.1109/TNSRE.2024.3438610.

29. Lee H, Choi YR, Lee HK, et al. Explainable vision transformer for automatic visual sleep staging on multimodal PSG signals. npj Digital Medicine. 2025;8(1). 10.1038/s41746-024-01378-0.

30. Zaschke P, Maurer MC, Hempel P, Hauschild AC, Rodenbeck A, Spicher N. A somnologist’s guide to explainable deep neural networks for sleep scoring. Somnologie. 2025;29(2):85–92. 10.1007/s11818-025-00504-8.

31. Van Der Donckt J, Van Der Donckt J, Deprost E, et al. Do not sleep on traditional machine learning: Simple and interpretable techniques are competitive to deep learning for sleep scoring. Biomedical Signal Processing and Control. 2023;81:104429. 10.1016/j.bspc.2022.104429.

32. Espiritu H, Metsis V. Automated Detection of Sleep Disorder-Related Events from Polysomnographic Data. In: 2015 International Conference on Healthcare Informatics;2015. p. 562–9. 10.1109/ICHI.2015.105.

33. Ugur TK, Erdamar A. An efficient automatic arousals detection algorithm in single channel EEG. Computer Methods and Programs in Biomedicine. 2019;173:131–8. 10.1016/j.cmpb.2019.03.013.

34. Liu Y, Liu H, Yang B. Automatic Sleep Arousals Detection From Polysomnography Using Multi-Convolution Neural Network and Random Forest. IEEE Access. 2020;8:176343–50. 10.1109/ACCESS.2020.3026814.

35. Rohan R, Kumari LVR. Classification of Sleep Apneas using Decision Tree Classifier. In: 2021 5th International Conference on Intelligent Computing and Control Systems (ICICCS);2021. p. 1310–6. 10.1109/ICICCS51141.2021.9432197.

36. Chambon S, Thorey V, Arnal PJ, Mignot E, Gramfort A. DOSED: A deep learning approach to detect multiple sleep micro-events in EEG signal. Journal of Neuroscience Methods. 2019;321:64–78. 10.1016/j.jneumeth.2019.03.017.

37. Brink-Kjaer A, Olesen AN, Peppard PE, et al. Automatic detection of cortical arousals in sleep and their contribution to daytime sleepiness. Clinical Neurophysiology. 2020;131(6):1187–203. 10.1016/j.clinph.2020.02.027.

38. Li H, Guan Y. DeepSleep convolutional neural network allows accurate and fast detection of sleep arousal. Communications Biology. 2021;4(1). 10.1038/s42003-020-01542-8.

39. Nassi TE, Ganglberger W, Sun H, et al. Automated Scoring of Respiratory Events in Sleep With a Single Effort Belt and Deep Neural Networks. IEEE Transactions on Biomedical Engineering. 2022;69(6):2094–104. 10.1109/TBME.2021.3136753.

40. Zahid AN, Jennum P, Mignot E, Sorensen HBD. MSED: A Multi-Modal Sleep Event Detection Model for Clinical Sleep Analysis. IEEE Transactions on Biomedical Engineering. 2023;70(9):2508–18. 10.1109/TBME.2023.3252368.

41. Nasiri S, Ganglberger W, Nassi T, et al. CAISR: achieving human-level performance in automated sleep analysis across all clinical sleep metrics. Sleep. 2025;48(8):zsaf134. 10.1093/sleep/zsaf134.

42. Rechtschaffen A, Kales A. A Manual of Standardized Terminology, Techniques and Scoring System for Sleep Stages of Human Subjects. Public Health Service. Washington DC: US Government Printing Office;1968.

43. Imtiaz SA, Rodriguez-Villegas E. Recommendations for performance assessment of automatic sleep staging algorithms. In: 2014 36th Annual International Conference of the IEEE Engineering in Medicine and Biology Society;2014. p. 5044-7. 10.1109/EMBC.2014.6944758.

44. Perslev M, Darkner S, Kempfner L, Nikolic M, Jennum PJ, Igel C. U-Sleep: resilient high-frequency sleep staging. npj Digital Medicine. 2021;4(1). 10.1038/s41746-021-00440-5.

45. Thorey V, Hernandez AB, Arnal PJ, During EH. AI vs Humans for the diagnosis of sleep apnea. In: 2019 41st Annual International Conference of the IEEE Engineering in Medicine and Biology Society (EMBC);2019. p. 1596–600. doi.org/10.1109/EMBC.2019.8856877.

46. Stephansen JB, Olesen AN, Olsen M, et al. Neural network analysis of sleep stages enables efficient diagnosis of narcolepsy. Nature Communications. 2018;9(1). 10.1038/s41467-018-07229-3.

47. Bakker JP, Ross M, Cerny A, et al. Scoring sleep with artificial intelligence enables quantification of sleep stage ambiguity: hypnodensity based on multiple expert scorers and auto-scoring. Sleep. 2022;46(2):zsac154. 10.1093/sleep/zsac154.

48. Anderer P, Ross M, Cerny A, Vasko R, Shaw E, Fonseca P. Overview of the hypnodensity approach to scoring sleep for polysomnography and home sleep testing. Frontiers in Sleep. 2023;Volume 2 - 2023. 10.3389/frsle.2023.1163477.

49. Friedman JH. Greedy Function Approximation: A Gradient Boosting Machine. The Annals of Statistics. 2001;29(5):1189–232. Available from: http://www.jstor.org/stable/2699986.

50. of Sleep Medicine AA. The AASM Manual for the Scoring of Sleep and Associated Events: Rules, Terminology and Technical Specifications. Version 2.6 ed. Darien, IL: American Academy of Sleep Medicine;2020.

51. Ke G, Meng Q, Finley T, et al. LightGBM: a highly efficient gradient boosting decision tree. In: Proceedings of the 31st International Conference on Neural Information Processing Systems. NIPS’17. Red Hook, NY, USA: Curran Associates Inc.;2017. p. 3149–57. 10.5555/3294996.3295074.

52. Welch P. The use of fast Fourier transform for the estimation of power spectra: A method based on time averaging over short, modified periodograms. IEEE Transactions on Audio and Electroacoustics. 1967;15(2):70–3. 10.1109/TAU.1967.1161901.

53. Harrison SJ, Bianchi S, Heinzle J, Stephan KE, Iglesias S, Kasper L. A Hilbert-based method for processing respiratory timeseries. NeuroImage. 2021;230:117787. 10.1016/j.neuroimage.2021.117787.

54. Martin Bland J, Altman D. STATISTICAL METHODS FOR ASSESSING AGREEMENT BETWEEN TWO METHODS OF CLINICAL MEASUREMENT. The Lancet. 1986;327(8476):307–10. Originally published as Volume 1, Issue 8476. 10.1016/S0140-6736(86)90837-8.

55. Redmon J, Divvala S, Girshick R, Farhadi A. You Only Look Once: Unified, Real-Time Object Detection. In: 2016 IEEE Conference on Computer Vision and Pattern Recognition (CVPR);2016. p. 779–88. 10.1109/CVPR.2016.91.

56. Guillot A, Sauvet F, During EH, Thorey V. Dreem Open Datasets: Multi-Scored Sleep Datasets to Compare Human and Automated Sleep Staging. IEEE Transactions on Neural Systems and Rehabilitation Engineering. 2020;28(9):1955–65. 10.1109/TNSRE.2020.3011181.

57. Rudin C. Stop explaining black box machine learning models for high stakes decisions and use interpretable models instead. Nature Machine Intelligence. 2019;1(5):206–15. 10.1038/s42256-019-0048-x.

58. Beam AL, Kohane IS. Big Data and Machine Learning in Health Care. JAMA. 2018;319(13):1317–8. 10.1001/jama.2017.18391.

59. Maher E, Epstein LJ. Artifacts and Troubleshooting. Sleep Medicine Clinics. 2009;4(3):421–434. 10.1016/j.jsmc.2009.04.010.

60. Rasquinha RJ, Moszczynski AJ, Murray BJ. A Modern Artifact in the Sleep Laboratory. Journal of Clinical Sleep Medicine. 2012;08(02):225–6. 10.5664/jcsm.1792.

61. Rinkevičius M, Charlton PH, Bailón R, Marozas V. Influence of Photoplethysmogram Signal Quality on Pulse Arrival Time during Polysomnography. Sensors. 2023;23(4). 10.3390/s23042220.

62. Zafar S, Ayappa I, Norman RG, Krieger AC, Walsleben JA, Rapoport DM. Choice of Oximeter Affects Apnea-Hypopnea Index. Chest. 2005;127(1):80–88. 10.1378/chest.127.1.80.

63. Liao YS, Wu MC, Li CX, Lin WK, Lin CY, Liang SF. Polysomnography scoring–related training and quantitative assessment for improving interscorer agreement. Journal of Clinical Sleep Medicine. 2024;20(2):271–8. 10.5664/jcsm.10852.

64. Penzel T, Zhang X, Fietze I. Inter-scorer Reliability between Sleep Centers Can Teach Us What to Improve in the Scoring Rules. Journal of Clinical Sleep Medicine. 2013;09(01):89–91. 10.5664/jcsm.2352.

65. Deng S, Zhang X, Zhang Y, et al. Interrater agreement between American and Chinese sleep centers according to the 2014 AASM standard. Sleep and Breathing. 2019;23(2):719–728. 10.1007/s11325-019-01801-x.

66. Nikkonen S, Somaskandhan P, Korkalainen H, et al. Multicentre sleep-stage scoring agreement in the Sleep Revolution project. Journal of Sleep Research. 2024;33(1):e13956. 10.1111/jsr.13956.

67. Dolek T, Akpinar D, Bek S, Cantas Turkis F, Kutlu G. A comparative analysis of automatic and manual scoring methods in polysomnography. Sleep. 2025;48(10):zsaf137. 10.1093/sleep/zsaf137.

68. Penzel T, Salanitro M. Emerging challenges in the transition from manual to automated sleep scoring. Sleep. 2025;48(10):zsaf202. 10.1093/sleep/zsaf202.

69. Feiner JR, Severinghaus JW, Bickler PE. Dark Skin Decreases the Accuracy of Pulse Oximeters at Low Oxygen Saturation: The Effects of Oximeter Probe Type and Gender. Anesthesia & Analgesia. 2007;105(6):S18–S23. 10.1213/01.ane.0000285988.35174.d9.

70. Sjoding MW, Dickson RP, Iwashyna TJ, Gay SE, Valley TS. Racial Bias in Pulse Oximetry Measurement. New England Journal of Medicine. 2020;383(25):2477–2478. 10.1056/nejmc2029240.

71. Al-Halawani R, Charlton PH, Qassem M, Kyriacou PA. A review of the effect of skin pigmentation on pulse oximeter accuracy. Physiological Measurement. 2023;44(5):05TR01. 10.1088/1361-6579/acd51a.

72. Javaheri S, Barbe F, Campos-Rodriguez F, et al. Sleep Apnea: Types, Mechanisms, and Clinical Cardiovascular Consequences. Journal of the American College of Cardiology. 2017;69(7):841–858. 10.1016/j.jacc.2016.11.069.

73. Hetland A, Vistnes M, Haugaa KH, Liland KH, Olseng M, Edvardsen T. Obstructive sleep apnea versus central sleep apnea: prognosis in systolic heart failure. Cardiovascular Diagnosis and Therapy. 2020;10(3). 10.21037/cdt.2020.03.02.

74. Ferri R, Koo BB, Picchietti DL, Fulda S. Periodic leg movements during sleep: phenotype, neurophysiology, and clinical significance. Sleep Medicine. 2017;31:29–38. Advances in Scientific Understanding of the Restless Legs Syndrome (RLS) (aka: Willis Ekbom Disease (WED)). 10.1016/j.sleep.2016.05.014.

75. Rathnayake SI, Abeyratne UR, Hukins C, Duce B. Modelling of polysomnographic respiratory measurements for artefact detection and signal restoration. Physiological Measurement. 2008;29(9):999. 10.1088/0967-3334/29/9/001.

76. Saifutdinova E, Congedo M, Dudysova D, Lhotska L, Koprivova J, Gerla V. An Unsupervised Multichannel Artifact Detection Method for Sleep EEG Based on Riemannian Geometry. Sensors. 2019;19(3). 10.3390/s19030602.

77. Forouzanfar M, Baker FC, Colrain IM, Goldstone A, de Zambotti M. Automatic analysis of pre-ejection period during sleep using impedance cardiogram. Psychophysiology. 2019;56(7):e13355. 10.1111/psyp.13355.

78. Anandanadarajah N, Talukder A, Yeung D, et al. Detection of Movement and Lead-Popping Artifacts in Polysomnography EEG Data. Signals. 2024;5(4):690–704. 10.3390/signals5040038.

79. Choo BP, Mok Y, Oh HC, et al. Benchmarking performance of an automatic polysomnography scoring system in a population with suspected sleep disorders. Frontiers in Neurology. 2023;Volume 14 - 2023. 10.3389/fneur.2023.1123935.

80. Gunnarsdottir KM, Gamaldo C, Salas RM, et al. A novel sleep stage scoring system: Combining expert-based features with the generalized linear model. Journal of Sleep Research. 2020;29(5):e12991. 10.1111/jsr.12991.

81. Holm B, Jouan G, Hardarson E, et al. An optimized framework for processing multicentric polysomnographic data incorporating expert human oversight. Frontiers in Neuroinformatics. 2024;Volume 18 - 2024. 10.3389/fninf.2024.1379932.

82. Rusanen M, Jouan G, Huttunen R, et al. Retrospective validation of automatic sleep analysis with grey areas model for human-in-the-loop scoring approach. Journal of Sleep Research. 2025;34(3):e14362. 10.1111/jsr.14362.

83. Lingren T, Deleger L, Molnar K, et al. Evaluating the impact of pre-annotation on annotation speed and potential bias: natural language processing gold standard development for clinical named entity recognition in clinical trial announcements. Journal of the American Medical Informatics Association. 2013;21(3):406–13. 10.1136/amiajnl-2013-001837.

84. Dai W, Yang Q, Xue GR, Yu Y. Boosting for transfer learning. In: Proceedings of the 24th International Conference on Machine Learning. ICML ’07. New York, NY, USA: Association for Computing Machinery;2007. p. 193–200. 10.1145/1273496.1273521.

85. Pan SJ, Yang Q. A Survey on Transfer Learning. IEEE Transactions on Knowledge and Data Engineering. 2010;22(10):1345–59. 10.1109/TKDE.2009.191.

86. Sun H, Jia J, Goparaju B, et al. Large-Scale Automated Sleep Staging. Sleep. 2017;40(10):zsx139. 10.1093/sleep/zsx139.

87. Leger D, Mutti C, Rouen A, Parrino L. Polysomnography in Transition: Reassessing Its Role in the Future of Sleep Medicine. Journal of Sleep Research. 2025;34(6):e70217. 10.1111/jsr.70217.

88. Li Q, Wen Z, Wu Z, et al. A Survey on Federated Learning Systems: Vision, Hype and Reality for Data Privacy and Protection. IEEE Transactions on Knowledge and Data Engineering. 2023;35(4):3347–66. 10.1109/TKDE.2021.3124599.

89. Rachakonda S, Moorthy S, Jain A, et al. Privacy Enhancing and Scalable Federated Learning to Accelerate AI Implementation in Cross-Silo and IoMT Environments. IEEE Journal of Biomedical and Health Informatics. 2023;27(2):744–55. 10.1109/JBHI.2022.3185418.

90. Salanitro M, van Acht V, Nijssen S, Randerath J, Fico G, Penzel T. Moving from Manual to Automated Sleep Staging with Federated Learning. In: 2025 47th Annual International Conference of the IEEE Engineering in Medicine and Biology Society (EMBC);2025. p. 1–4. 10.1109/EMBC58623.2025.11253097.

91. Fiorillo L, Puiatti A, Papandrea M, et al. Automated sleep scoring: A review of the latest approaches. Sleep Medicine Reviews. 2019;48:101204. 10.1016/j.smrv.2019.07.007.

